# Combined use of absolutely quantitated cyclin D1 and Ki67 protein levels to improve prognosis of Luminal-like breast cancer

**DOI:** 10.1101/2020.04.15.20066993

**Authors:** Junmei Hao, Wenfeng Zhang, Yan Lv, Jiarui Zou, Yunyun Zhang, Jiahong Lv, Shuishan Xie, Cuiping Zhang, Jiandi Zhang, Fangrong Tang

## Abstract

**Purpose:** Both Ki67 and cyclin D1 are routinely used protein biomarkers of cell proliferation for breast cancer patients. Ki67 is used to differentiate Luminal A-like from Luminal-B like subtype in surrogate assay. These two proliferative factors are investigated in this retrospective study to evaluate their prognostic role on the overall survival (OS) of Luminal-like breast cancer patients.

**Method:** The cyclin D1 protein level was measured absolutely and quantitatively using Quantitative Dot Blot (QDB) method in 143 Luminal-like FFPE breast cancer specimens. An optimized cutoff at 0.71 μmole/g was identified and used to separate these specimens into cyclin D1 high and low groups alone, or in combination with Ki67, for overall survival (OS) analyses of these patients.

**Results:** Cyclin D1 was found to be an independent prognostic factor from Ki67 in univariate and multivariate analysis. When both biomarkers were used to separate these Luminal-like specimens, the group with low expression of both biomarkers (n=52) had significantly improved 10 year survival probability at 94%, while the one with high expression of both markers (n=34) were at 41% based on Kaplan-Meier survival analysis of OS (Log rank test p<0.0001).

**Conclusion:** We demonstrated cyclin D1 as an independent prognostic protein biomarker from Ki67 for Luminal-like breast cancers. The combined usage of cyclin D1 and Ki67 significantly improved the prognosis over current prevailing surrogate assay. We propose to incorporate cyclin D1 in surrogate assay to improve prognosis for Luminal-like breast cancer patients in future clinical practice.

## Introduction

The Luminal-like breast cancer, featured by overexpression of estrogen receptor (ER), is defined in surrogate assay as tumors with ≥1% positive staining of ER with immunohistochemistry (IHC) in daily clinical practice[1–4]. This group of tumors are further separated into Luminal A-like and Luminal B-like subtypes based on a protein biomarker of cell proliferation, Ki67. More specifically, based on IHC and/or Fluorescence in situ hybridization (FISH) analyses, those tumors with Ki67<14%, Progesterone receptor (PR) >20%, and Her2 negative (Her2-) are defined as Luminal A-like subtype with better prognosis. The rest of the Luminal-like tumors are classified as Luminal B-like subtype, including both B_1_ (ER+, Her2-, Ki67>14% or PR<20%) and B_2_ (ER+, Her2+) subgroups.

Cyclin D1 is another commonly used protein biomarker of cell proliferation in routine clinical practice. While the exact biological role of Ki67 remains unclear until now[5], cyclin D1 is known as the key regulator of cell cycle progression. Overexpression of cyclin D1 drives the cells from G1 to S phase in mitosis[6]. In this study, we set out to investigate if there were any prognostic differences between these two protein biomarkers of cellular proliferation. More importantly, if we can improve prognosis using both protein biomarkers simultaneously.

In a previous study, we measured Ki67 protein levels absolutely and quantitatively in 143 Luminal-like Formalin Fixed Paraffin Embedded (FFPE) specimens using our independently developed Quantitative Dot Blot (QDB) method[7]. When replacing Ki67 score from IHC analysis with our absolutely quantitated Ki67 levels, we found the prognosis of surrogate assay was improved significantly for OS of Luminal-like tumors[7].

Along this line of thinking, in this study, the cyclin D1 protein levels were also measured quantitatively and absolutely using QDB method in the same 143 Luminal-like FFPE specimens reported in the previous study, and we used these results to demonstrate that cyclin D1 was an independent prognostic factor from Ki67. The combined usage of these two protein biomarkers achieved significantly better prognosis than that of surrogate assay for Luminal-like tumors.

## Materials and Methods

### Human subjects and human cell lines

A total of 143 Formalin Fixed Paraffin Embedded (FFPE) breast cancer tissues in 2×15 µm slices treated between 2008 and 2013 at Yantai Affiliated Hospital of Binzhou Medical University (Yantai, P. R. China) were collected sequentially. All the samples were obtained in accordance with the Declaration of Helsinki and were approved by the Medical Ethics Committee of Yantai Affiliated Hospital of Binzhou Medical University (Approval #: 20191127001), with an informed Consent Forms waiver for archived specimens.

MCF-7 cell line was purchased from the Cell Bank of Chinese Academy of Sciences (Shanghai, China) and cultured as provider’s instruction.

### General reagents

The general reagents for cell culture were purchased from Thermo Fisher Scientifics (Waltham, MA, USA). The chemicals used for protein expression were purchased from Takara Inc. (Dalian, China), and Nickel-His GraviTrap affinity column for protein purification was purchased from GE Healthcare. All the other chemicals were purchased from Sinopharm Chemicals (Beijing, P. R. China).

Rabbit anti-cyclin D1 monoclonal antibody (EP12) was purchased from ZSGB-BIO (Beijing, China). HRP labeled Donkey Anti-Rabbit IgG secondary antibody was purchased from Jackson Immunoresearch lab (Pike West Grove, PA, USA). Pierce BCA protein quantification kit was purchased from Thermo Fisher Scientific Inc. (Calsband, CA, USA). Recombinant human cyclin D1 protein was prepared in the house. QDB plate was manufactured by Quanticision Diagnostics Inc at RTP, NC, USA.

### Preparation of recombinant cyclin D1 protein

A DNA sequence corresponding to the 271-295AA of human cyclin D1 (NCBI #: NP_444284.1) was inserted into PET32a (+) expression vector and verified by DNA sequencing. The plasmid was expressed in BL21 (DE3) competent cells after induction of IPTG. Total bacterial protein lysate was extracted in binding buffer (20 mM sodium phosphate, 500 mM NaCI, 20 mM imidazole, PH 7.4) by ultrasonication. Recombinant cyclin D1 protein of bacterial lysate was purified by Ni-NTA column according to the instructions. Fractions containing cyclin D1 protein were pooled and concentrated on a Milipore ultrafiltration spin column. Protein concentration of cyclin D1 was determined using BCA protein quantification kit, and the purity was determined by SDS-PAGE before aliquots were stored at −80 °C.

### Preparation of FFPE tissues and cell lysates

Breast cancer FFPE specimens in 2×15 µm slices were de-paraffinized and solubilized with lysis buffer (50 mM HEPES, 137 mM NaCl, 5 mM EDTA, 1 mM MgCl, 10 mM Na_2_P_2_O_7_, 1% TritonX-100, 10% glycerol). MCF-7 cell pellet was collected and Formalin-fixed for 30min before solubilization with the same lysis buffer. The supernatants were collected after centrifugation for QDB analysis. Protein concentration was detected using BCA protein quantification kit.

### QDB analysis

The QDB process was described previously with minor modifications[8–11]. In brief, FFPE tissue lysates (about 0.35 μg/unit) and MCF-7 cell lysate (about 0.08 μg/unit) were applied onto the QDB plate at 2 μl/unit in triplicate. Serially diluted cyclin D1 recombinant protein was included in each plate as protein standard for calculation of the absolute cyclin D1 protein levels. The loaded QDB plate was dried in the air for 4h and then blocked with blocking buffer (4% non-fat milk in TBST) for 1h. The plate was inserted into 96-well microplate pre-filled with 100μl anti-cyclin D1 antibody (clone EP12, 1:500 in blocking buffer) and incubated overnight at 4°C. The plate was rinsed twice and washed 4×10 mins before it was incubated with a donkey anti-rabbit secondary antibody for another 4 hours at room temperature. The plate was then rinsed twice and washed 5×10 mins before being processed with ECL reagent and quantified with a Tecan Microplate reader. The chemiluminescence signals were converted into absolute cyclin D1 protein level in each sample using protein standard. All results were averages of three independent experiments, with each sample in triplicate.

MCF-7 cell lysate was used as internal control. The absolute cyclin D1 level of MCF-7 cell lysate was measured with QDB method and the results were documented. The overall experiment was judged as valid when the measured cyclin D1 level of MCF-7 cell lysate fell within ±20% of the documented value.

### Statistical analysis

All the statistical analyses were done using either R version 3.6.2 (http://www.r-project.org) or GraphPad Prism 7 (La Jolla, CA, USA). The results were reported as mean ± standard error of the mean (SEM). A p-value of <0.05 was defined as statistically significant. The correlation between cyclin D1 and Ki67 protein levels was analyzed with Spearman’s correlation coefficient analysis. Differences in the clinicopathologic features between patients assigned to C_l_K_l_ and C_h_K_h_ groups were examined using Chi-squared test.

The endpoint of overall survival analysis was defined as the time from breast surgery to death or last follow-up (April 1st, 2019). Survival data for patients who were still alive at the date of last follow-up were treated as censored. Cyclin D1 levels measured by QDB method were dichotomized for OS analysis, using optimized cutoff value determined by the “surv_cutpoint” function of the “surviminer” R package. A total of 143 Luminal-like patients in surrogate assay were grouped into low cyclin D1 group and high cyclin D1 group.

The relationship between Ki67 and/or cyclin D1 and OS of Luminal-like specimens was evaluated using the log-rank test and visualized by Kaplan-Meier survival curve analysis. Univariate Cox proportional hazard model fitted overall survival was employed for hazard ratio (HR) and corresponding 95% confidence intervals (CIs) estimation. Multivariable Cox model was used to examine the association between Ki67 and/or cyclin D1 and OS with adjusting for clinical factors, such as age, node status, tumor size, tumor grade, and type of treatment. The Schoenfeld residuals test is used to test the proportional Hazard assumption in Cox Model.

## Results

The clinicopathological characteristics of all 143 Luminal-like FFPE specimens were described in Table. 1. These specimens were collected sequentially from local hospital, as described in the Materials and Methods section.

**Table 1.**
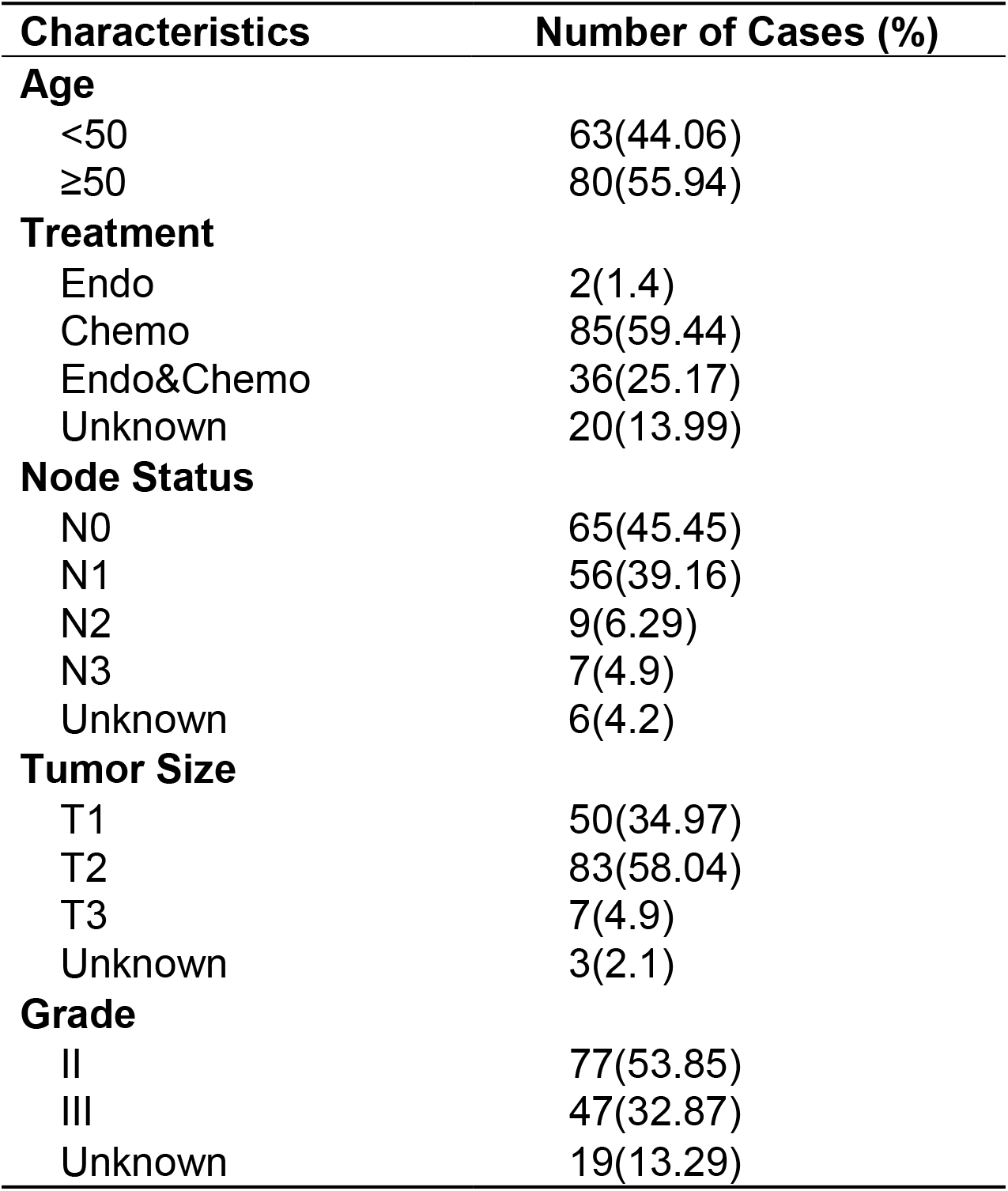
Clinicopathological Characteristics of 143 Luminal-like specimens

Total protein lysates were extracted from 2×15 μm FFPE tissue slices from each individual specimen. The linear ranges of both total tissue lysates and recombinant cyclin D1 protein were defined using QDB method (Supplemental Fig. 1). As shown in Fig. 1a, the distribution of cyclin D1 levels were from 0.01 to 5.63 μmole/g, with mean at 0.83±0.07 μmole/g. The expression levels of cyclin D1 were weakly correlated with those of Ki67 when analyzed using Spearman’s correlation analysis (ρ=0.29, p=0.0005, n=143).

**Figure 1.**
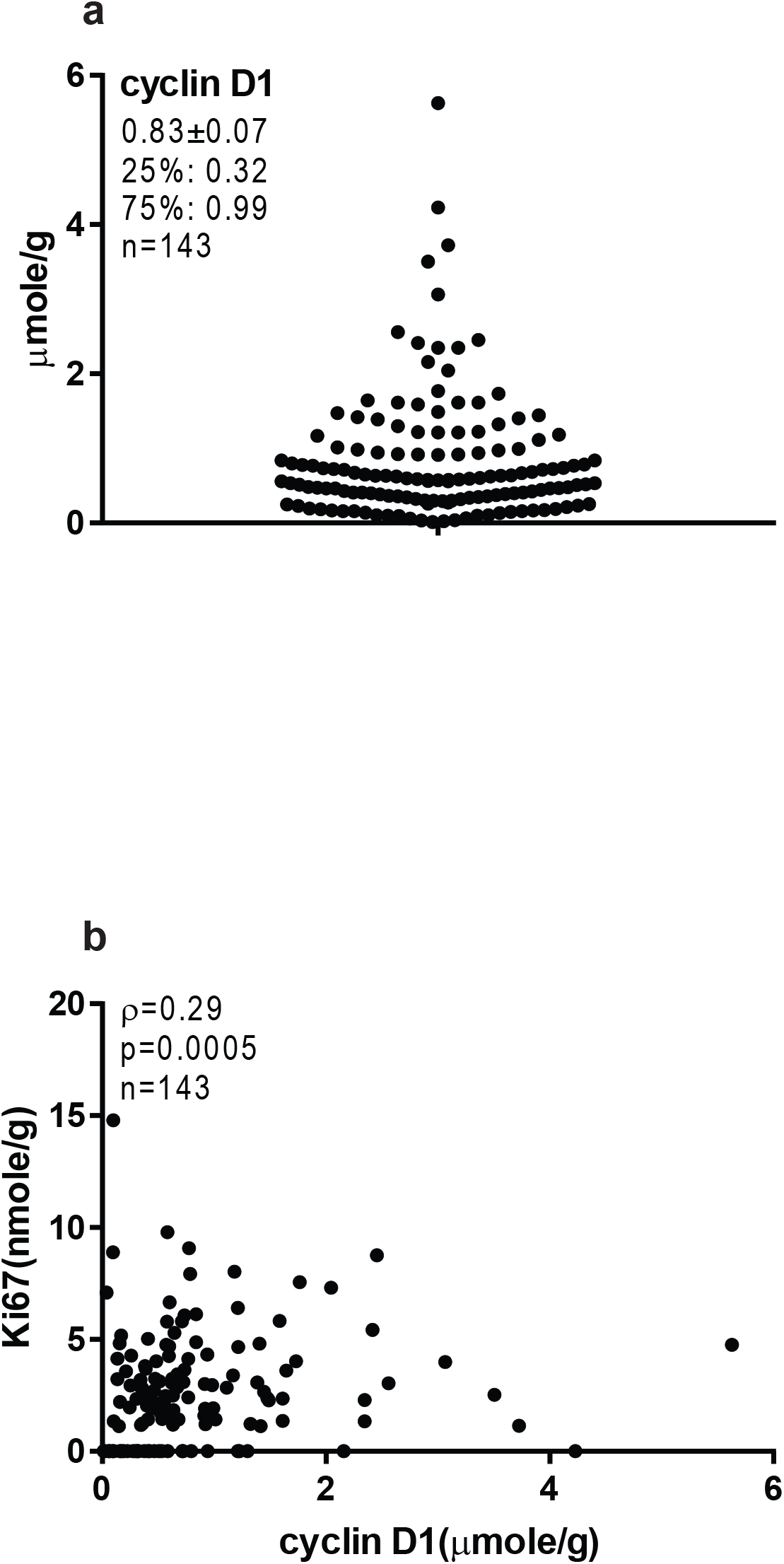
The cyclin D1 protein levels in 143 Luminal-like breast cancer FFPE tissues. A total of 143 Luminal-like breast cancer FFPE tissues in 2×15 μm slices were collected sequentially from local hospital, and the total protein lysates were prepared as described in Materials and Methods. MCF-7 cell lysate was used as internal control. FFPE tissue lysates (about 0.35 μg/unit) and cell lysate (about 0.08 μg/unit) were applied onto the QDB plate at 2 μl/unit in triplicate for QDB analysis using anti-cyclin D1 antibody. Serially diluted cyclin D1 recombinant protein were included in each plate as protein standard. All results were averages of three independent experiments, with each sample in triplicate. **(a)** cyclin D1 levels were from 0.01 to 5.63 μmole/g, with mean at 0.83 μmole/g and median at 0.58 μmole/g. **(b)** The correlation between cyclin D1 and Ki67 protein levels in 143 FFPE tissues was analyzed with Spearman’s correlation coefficient analysis using Graphpad software, ρ=0.29, p=0.0005.

The putative prognostic role of cyclin D1, alongside with Ki67, for OS of Luminal-like breast tumors was explored first with univariate survival analysis (Table. 2). Both cyclin D1 and Ki67 were found to be independent prognostic factor for the OS of the patients, with HR for Ki67 at 1.18 (95% CI: 1.05-1.31, p=0.0035) and for cyclin D1 at 1.47 (95% CI: 1.12-1.93, p=0.0056) respectively. In addition, other factors including node status and age were also found to be statistically significant, while the tumor size, treatment and tumor grade were found not to be significantly associated with OS of the patients.

**Table 2.**
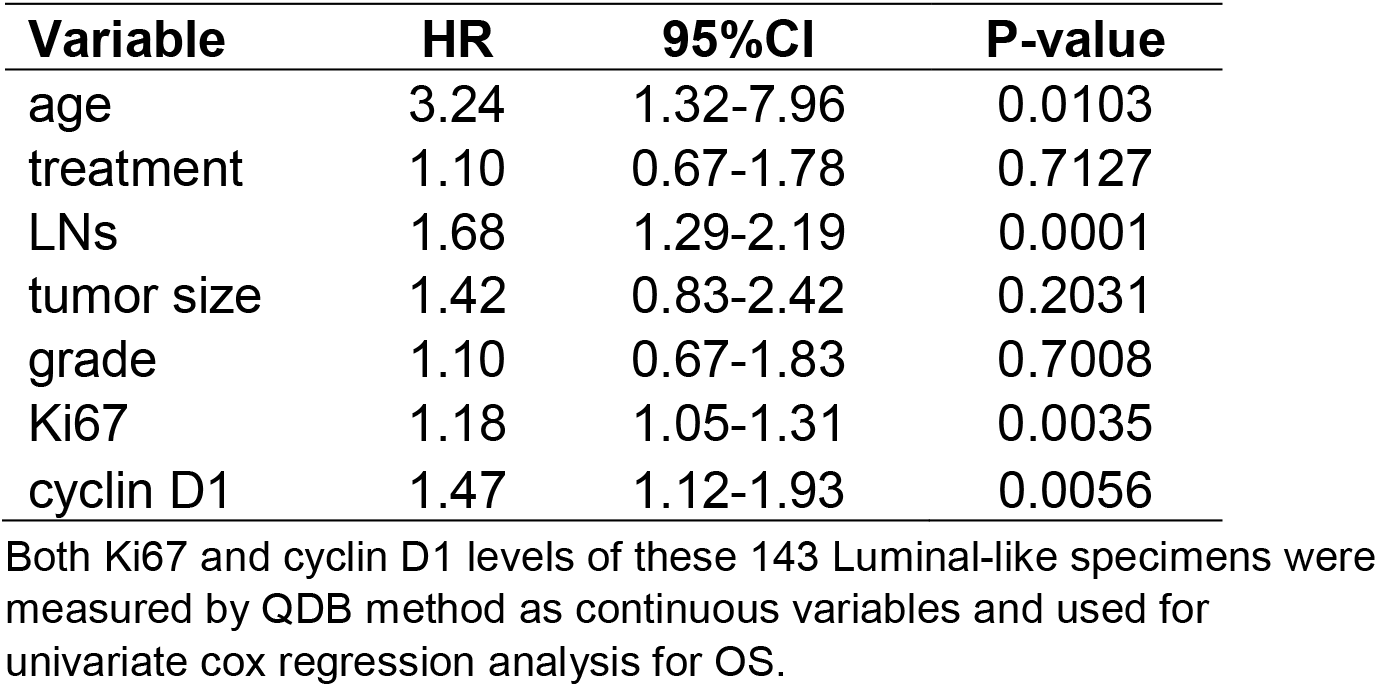
Univariate Cox regression analysis of Overall Survival (OS) of Ki67 and cyclin D1 protein levels

| **Variable** | **HR** | **95%CI** | **P-value** |
| --- | --- | --- | --- |
| age | 3.24 | 1.32-7.96 | 0.0103 |
| treatment | 1.10 | 0.67-1.78 | 0.7127 |
| LN's | 1.68 | 1.29-2.19 | 0.0001 |
| tumor size | 1.42 | 0.83-2.42 | 0.2031 |
| grade | 1.10 | 0.67-1.83 | 0.7008 |
| Ki67 | 1.18 | 1.05-1.31 | 0.0035 |
| cyclin D1 | 1.47 | 1.12-1.93 | 0.0056 |

Both Ki67 and cyclin D1 levels of these 143 Luminal-like specimens were measured by QDB method as continuous variables and used for univariate cox regression analysis for OS.

The prognostic roles of both cyclin D1 and Ki67 for OS were next analyzed with multivariate cox regression survival analysis. Unexpectedly, although both factors are biomarkers of cell proliferation, they were found to be independent prognostic factor from each other, with HR for Ki67 at 1.19 (95% CI: 1.04-1.37, p=0.0138) and for cyclin D1 at 1.45 (95% CI: 1.04-2.02, p=0.0291). Again, the age and node status were found to be independent prognostic factor in the same analysis.

The fact that cyclin D1 and Ki67 were independent prognostic factors from each other prompted us to investigate if we can improve the prognosis of Luminal-like tumors using both factors. Therefore, we tried first to identify the optimized cyclin D1 cutoff value to separate Luminal-like specimens for OS analysis. This value was determined by the “surv_cutpoint” function of the “surviminer” R package at 0.71 μmole/g and used to separate these Luminal-like specimens into cyclin D1 high (C_h_) and low (C_l_) groups. The OS analysis of the resulted C_h_ and C_l_ group was done using Kaplan-Meier survival analysis (Fig. 2). We found C_l_ group have 10-year survival probability at 89%, in comparison to the 57% for C_h_ group, with p<0.0001 from Log Rank test. As an comparison, the prognosis from adjusted surrogate assay, where the Luminal-like specimens were subtyped based on absolutely quantitated Ki67 protein levels, had the 10-year survival probability at 91% for Luminal A-like subtype, and 63% for Luminal B-like subtype, with p=0.00052[7]. For current prevailing surrogate assay, the 10-year survival probability was at 88% for Luminal A-like subtype, and 68% for Luminal B-like subtype with p=0.031 from Log Rank test using the same specimens.

**Figure 2.**
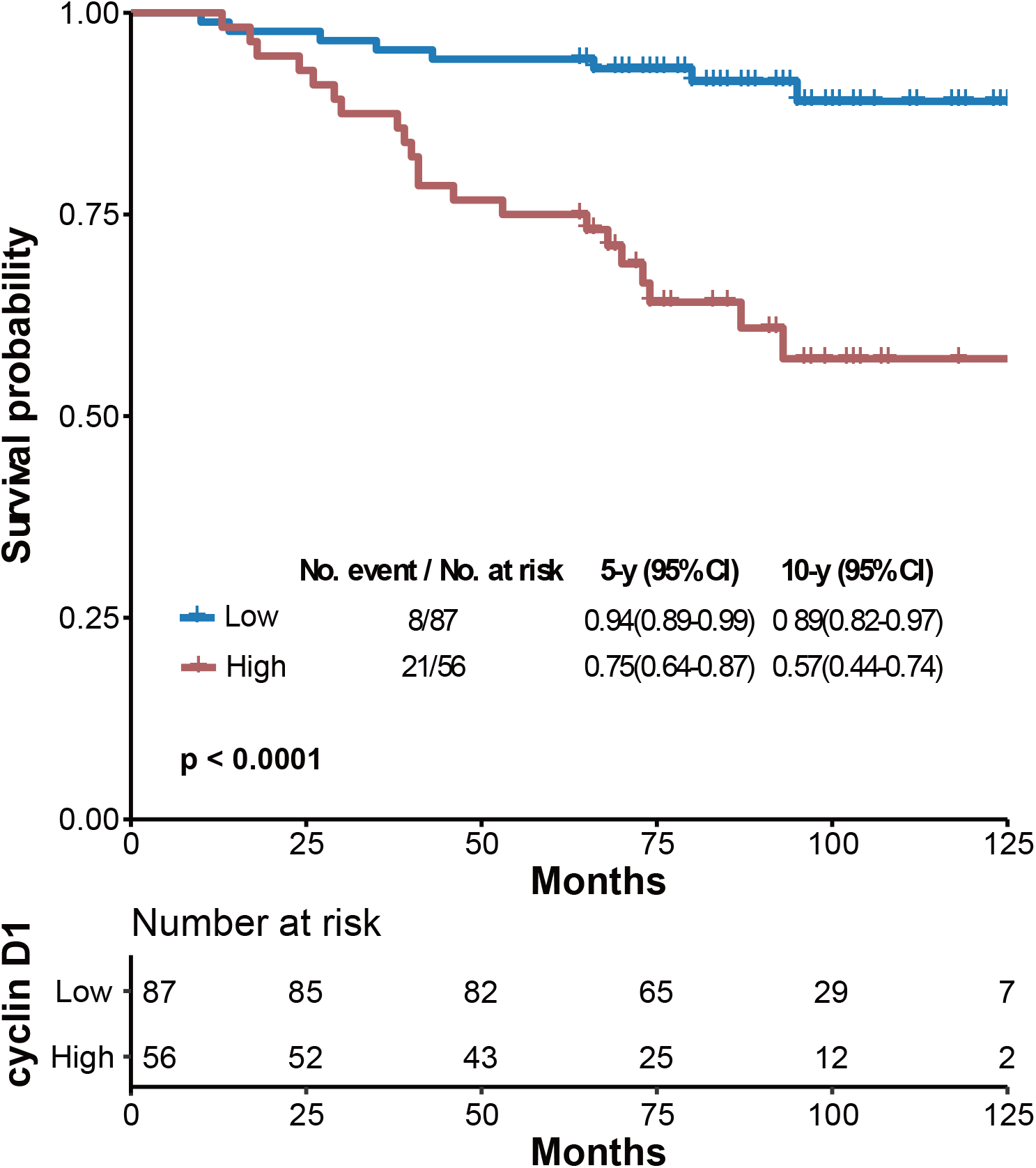
Kaplan-Meier curves for overall survival (OS) of Luminal-like specimens divided into cyclin D1 low (C_l_) and high groups (C_h_) by the protein levels of Cyclin D1, using 0.71 μmole/g as cutoff. Optimal cutoff value of cyclin D1 level was determined at 0.71 μmole/g by the “surv_cutpoint” function of the “surviminer” R package. The 5-year and 10-year survival probabilities, and the p values from Log Rank test were provided in the figure. CI, confidence interval; p, log-rank p-value.

These Luminal-like specimens were also separated into Ki67 high (K_h_) and Ki67 low (K_l_) group using the 2.31 nmole/g we defined in the previous study[7]. These two cutoffs were used together to further separate the Luminal-like specimens into four groups, with C_h_K_h_ for specimens with the expression levels of both biomarkers above the respective cutoffs, C_l_K_l_ for those below the respective cutoffs. C_h_K_l_ refers to specimens with only cyclin D1 level above the corresponding cutoff, while the C_l_K_h_ refers to those with only Ki67 level above the corresponding cutoff. The OS of these four groups were analyzed using Kaplan-Meier survival analysis in Fig. 3. We found the C_l_K_l_ group (n=52) had significantly improved 10-year survival probability at 94% (95% CI: 0.86-1), while C_h_K_h_ group (n=34) had the worst 10-year survival probability at 41% (95% CI: 0.24-0.69). The C_h_K_l_ (n=22) and C_l_K_h_ (n=35) group had similar 10-year survival probability at 77% (95% CI: 0.61-0.97) and 83% (95% CI: 0.71-0.96) respectively. The p value from Log rank test was <0.0001. It should be emphasized that in this simple categorization, the potential impact of PR is not considered. In fact, the two deaths in C_l_K_l_ group were all found to be associated with low PR expression (unpublished data).

**Figure 3.**
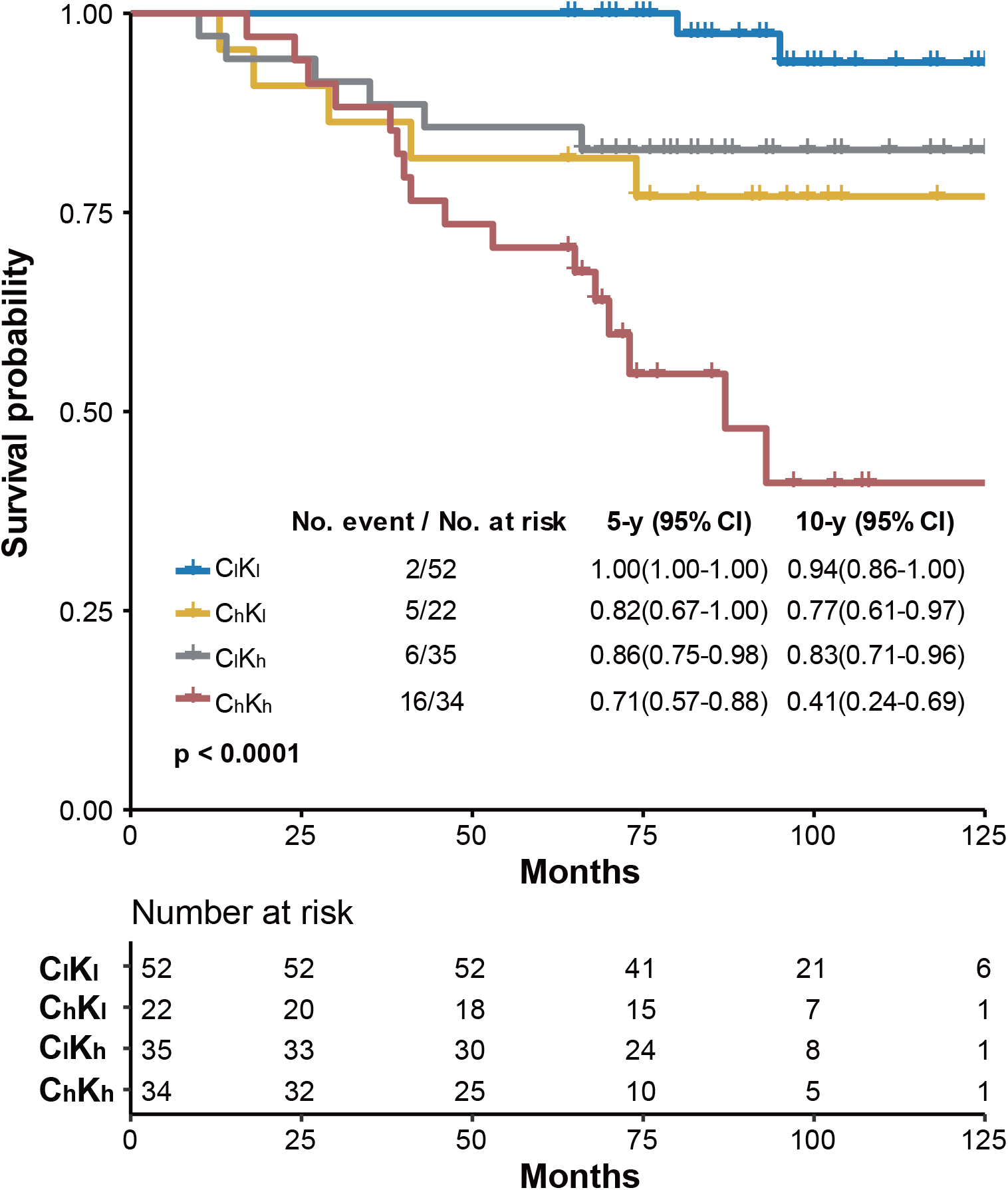
Kaplan-Meier curves for overall survival (OS) of Luminal-like specimens grouped by both the Ki67 and Cyclin D1 protein levels. The specimens were separated into four groups using Cyclin D1 at 0.71 μmole/g and Ki67 at 2.31 nmole/g as cutoffs respectively. C_l_K_l_ stands for specimens with the protein levels of both biomarkers below the respective cutoffs, C_l_K_h_ for those with only Cyclin D1 level below the recommended cutoff, C_h_K_l_ for those with only Ki67 levels below the recommended cutoff, and C_h_K_h_ for those with both biomarkers above the respective cutoffs. The 5-year and 10-year survival probabilities, and the p values from Log Rank test were provided in the figure.

The clinicopathological parameters from both C_l_K_l_ group with the best prognosis, and C_h_K_h_ group with the worst prognosis, were also compared in Supplemental table. 1. We found that these two groups were similar at age, treatment, and node status. Consistent with the prior knowledge, there were more patients with T1 stage in tumor size, and grade II in histological grade in C_l_K_l_ group than those of C_h_K_h_ group. Yet, none of them reached statistically significance due to the small sample size.

## Discussion

In this study, the cyclin D1 levels were absolutely and quantitatively measured using QDB method in 143 Luminal-like FFPE specimens. We found that cyclin D1 was an independent prognostic factor from Ki67 for OS of Luminal-like patients. More importantly, we found the combined use of Ki67 and cyclin D1 significantly improved the prognosis of OS of Luminal-like breast cancer.

As shown in table 4, when both biomarkers were used, we identified C_l_K_l_ group with the best prognosis (n=52), with 10-year survival probability at 94%, in comparison to the Luminal A-like subtype at 91% for adjusted surrogate assay (n=69), and 88% for surrogate assay (n=61); and C_h_K_h_ group with the worst prognosis (n=34) with 10-year survival probability at 41%, in comparison to 63% for Luminal B-like subtype from adjusted surrogate assay (n=74) and 68% from surrogate assay (n=82) [7]. In addition, in C_h_K_h_ group, the percentage of deceased patients were 47.05% (16/34), in comparison to 31.08% (23/74) in Luminal B-like subtype from adjusted surrogate assay and 26.83% (22/82) from surrogate assay.

**Table 3.** Multivariate Cox regression analysis of Overall Survival (OS) of Ki67 and cyclin D1 protein levels adjusted for clinical variables

| **Variable** | **HR** | **95%CI** | **P-value** |
| --- | --- | --- | --- |
| Age | 4.11 | 1.61-10.47 | 0.0030 |
| Treatment | 0.90 | 0.54-1.51 | 0.7030 |
| Node Status | 1.72 | 1.26-2.34 | 0.0007 |
| Tumor Size | 1.35 | 0.67-2.75 | 0.4028 |
| Grade | 0.87 | 0.45-1.68 | 0.6825 |
| Ki67 | 1.19 | 1.04-1.37 | 0.0138 |
| cyclin D1 | 1.45 | 1.04-2.02 | 0.0291 |

Multivariate survival analysis was performed using the Cox proportional hazards model adjusted for age, treatment, node status, tumor size and tumor grade.

**Table 4.**
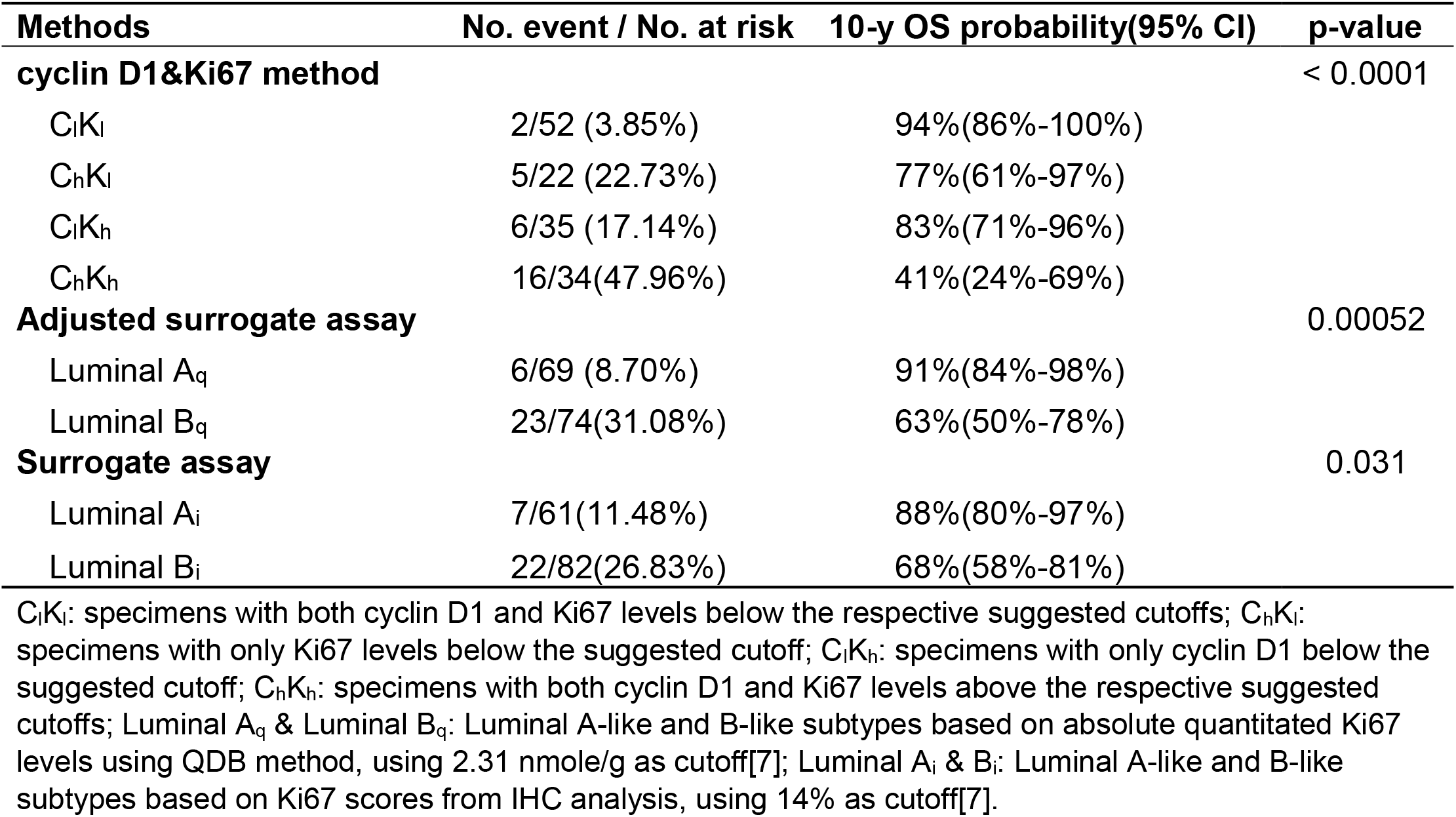
Performance of three subtyping methods

Although there are extensive studies about the prognostic role of cyclin D1 among breast cancer patients, the conclusion remains controversial, even among patients of ER+. All these results were well summarized in Ahlin et al and several other studies[6, 12, 13]. We suspect these studies were seriously limited by the method used in these studies. For example, IHC is used primary for assessment of cyclin D1 protein levels in breast cancer specimens[6]. Yet, this method is known to be associated with inconsistency and subjectivity[14]. By using QDB method in this study, these issues were well circumvented, and we were able to show a strong correlation of overexpression of cyclin D1 with reduced survival of ER+ patients, supported by both the univariate and multivariate cox regression survival analyses.

More importantly, we demonstrated for the first time that cyclin D1 is an independent prognostic biomarker from Ki67. Thus, the combined use of these two biomarkers in daily clinical practice should significantly improve the prognosis of Luminal-like patients. Although the role of cyclin D1 in basal-like tumors remains to be explored, considering that around 60∼70% tumors are Luminal-like tumors[3], cyclin D1 is highly recommended to be included in the routine subtyping practice for breast cancer patients.

In this study, we also separate all the Luminal-like specimens into four groups of C_h_K_h,,_ C_h_K_l_, C_l_K_h_ and C_l_K_l_ simply using our proposed cutoffs at 0.71 μmole/g for cyclin D1 and 2.31 nmole/g for Ki67, without considering the potential impact of PR and Her2 in the analysis. Even under this crude classification, we still provided a better prognosis than that from the current available surrogate assay.

It should be emphasized that this study is by no means to dismiss the prognostic role of PR. Rather, this factor is yet to be incorporated in current prognostic method. In fact, consistent with the protective role of PR in surrogate assay, the only two deaths in the C_l_K_l_ group in our study are with low expression of PR (unpublished data). Clearly, with more specimens available, a better algorithm will be developed to incorporate PR, cyclin D1 and Ki67 in the prognosis evaluation of Luminal-like patients.

It should also be pointed out that this is only a preliminary study, with merely 143 Luminal-like FFPE specimens available for analysis. The analysis was also only limited to the OS of these patients. More retrospective and prospective studies are urgently needed to verify our conclusion, so that patients worldwide can benefit from this finding sooner. The prognostic effect of cyclin D1 on the Disease-free survival (DFS) should also be investigated in the future.

In conclusion, by measuring cyclin D1 protein levels absolutely and quantitatively in 143 Luminal-like FFPE specimens, we demonstrated that overexpression of cyclin D1 was negatively associated with the survival of Luminal-like patients, and the combined usage of cyclin D1 and Ki67 significantly improve the prognosis of Luminal-like patients. We proposed to use cyclin D1 together with Ki67 protein biomarkers routinely in daily clinical practice to aid subtyping of breast cancer patients.

## Data Availability

All the data are available upon reasonable request by writing to

## Author contributions

JH, JZou, SX & CZ provided clinical samples & JH supervised all the clinical studies; WZ, YL, YZ, JL, & FT performed all the assays and data analysis; FT supervised all the assays; YL performed all the statistical analysis, JZhang designed & supervised the overall study and drafted the manuscript; WZ, JZhang & FT contributed to data interpretation and edited the manuscript.

---

**Supplemental fig 1.**
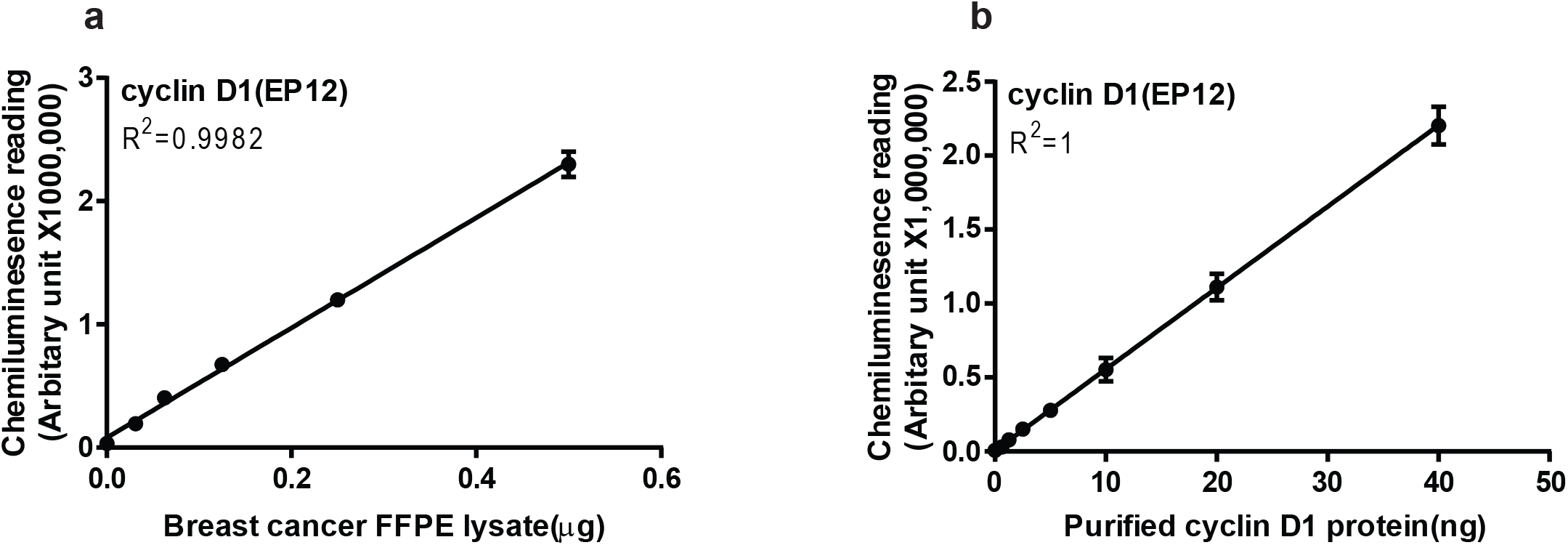
Defining the linear ranges of QDB analysis. **(a)** Defining the linear range of QDB method with breast cancer FFPE lysate. Human breast cancer FFPE specimens in 2×15 μm slices were provided sequentially by local hospital, and the pooled FFPE lysate was prepared by mixing in equal amount of 4 total tissue lysates with high IHC scores for cyclin D1. The pooled lysate was serially diluted as indicated in the figure and supplemented with IgG-Free BSA solution to allow for equal loading (about 0.5 μg/unit). The lysate was applied onto QDB plate at 2 μl/unit for QDB analysis using anti-cyclin D1 antibody (clone EP12). **(b)** Defining the linear range of QDB method with purified cyclin D1 protein. The cyclin D1 recombinant protein was serially diluted as indicated in the figure and supplemented with IgG-Free BSA solution. The diluted recombinant protein lysate was applied onto the QDB plate at 2 μl/unit for QDB analysis using anti-cyclin D1 antibody (clone EP12).

**Supplemental table 1.**
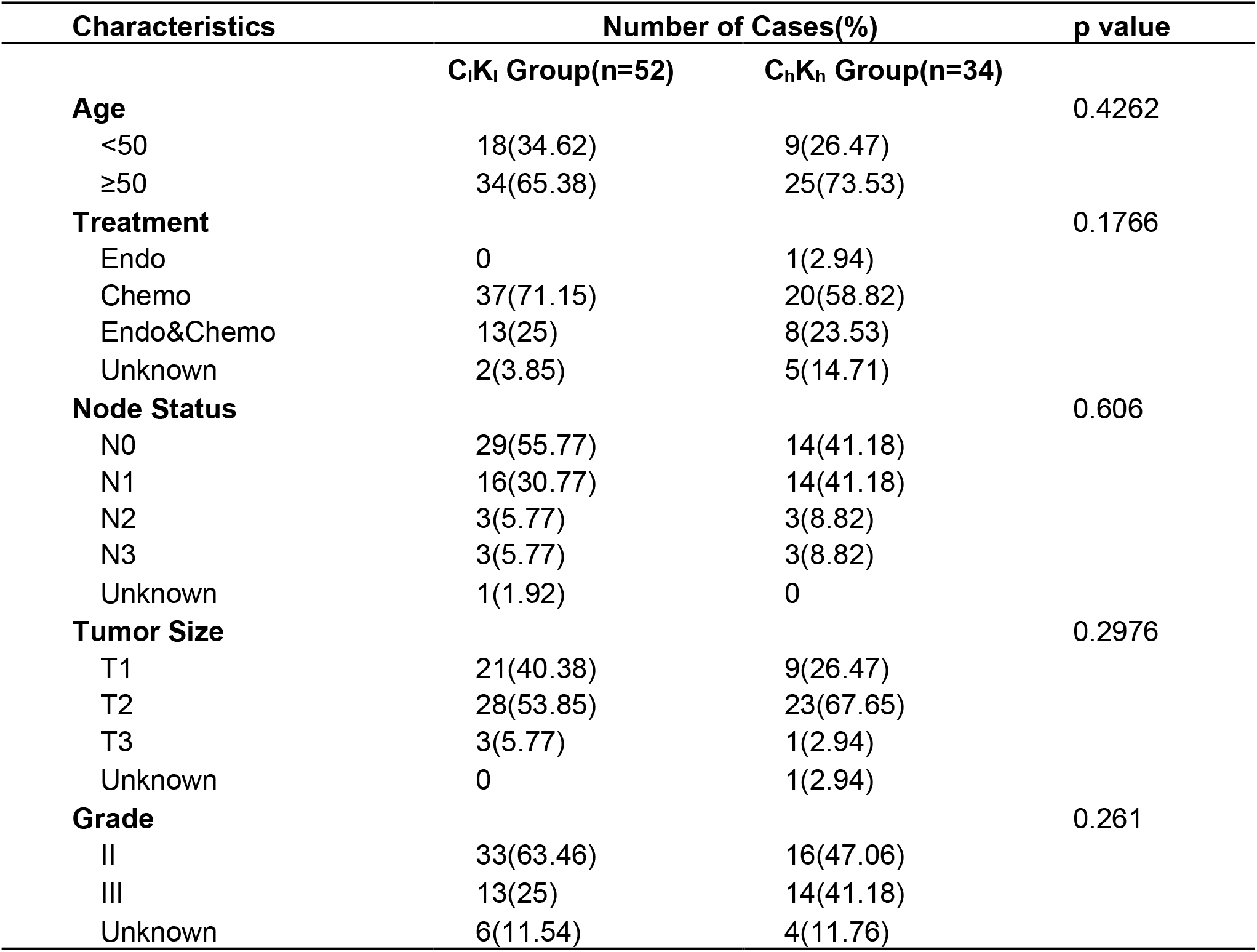
Clinicopathological characteristics of specimens in C_l_K_l_ and C_h_K_h_ groups

## References

1. Perou CM, Sørlie T, Eisen MB et al. Molecular portraits of human breast tumours. Nature 2000; 406(6797):747–752.

2. Goldhirsch A, Winer EP, Coates AS et al. Personalizing the treatment of women with early breast cancer: highlights of the St Gallen International Expert Consensus on the Primary Therapy of Early Breast Cancer 2013. Ann Oncol 2013; 24(9):2206–2223.

3. Yersal O, Barutca S. Biological subtypes of breast cancer: Prognostic and therapeutic implications. World J Clin Oncol 2014; 5(3):412–424.

4. Sørlie T, Perou CM, Tibshirani R et al. Gene expression patterns of breast carcinomas distinguish tumor subclasses with clinical implications. Proc. Natl. Acad. Sci. U.S.A. 2001; 98(19):10869–10874.

5. Juríková M, Danihel Ľ, Polák Š, Varga I. Ki67, PCNA, and MCM proteins: Markers of proliferation in the diagnosis of breast cancer. Acta Histochemica 2016; 118(5):544–552.

6. Ahlin C, Lundgren C, Embretsén-Varro E et al. High expression of cyclin D1 is associated to high proliferation rate and increased risk of mortality in women with ER-positive but not in ER-negative breast cancers. Breast Cancer Res Treat 2017; 164(3):667–678.

7. Hao J, Lv Y, Zou J et al. Improving prognosis of surrogate assay for breast cancer patients by absolute quantitation of Ki67 protein levels using Quantitative Dot Blot (QDB) method. medRxiv 2020:2020.03.11.20034439.

8. Tian G, Tang F, Yang C et al. Quantitative dot blot analysis (QDB), a versatile high throughput immunoblot method. Oncotarget 2017; 8(35):58553–58562.

9. Zhang W, Yu G, Zhang Y et al. Quantitative Dot Blot (QDB) as a universal platform for absolute quantification of tissue biomarkers. Analytical Biochemistry 2019; 576:42–47.

10. Yu G, Zhang W, Zhang Y et al. Developing a routine lab test for absolute quantification of Her2 in Formalin Fixed Paraffin Embedded (FFPE) breast cancer tissues using Quantitative Dot Blot (QDB) method. bioRxiv 2019:584615.

11. Qi X, Zhang Y, Zhang Y et al. High Throughput, Absolute Determination of the Content of a Selected Protein at Tissue Levels Using Quantitative Dot Blot Analysis (QDB). JoVE (Journal of Visualized Experiments) 2018; (138):e56885–e56885.

12. Lundberg A, Lindström LS, Harrell JC et al. Gene Expression Signatures and Immunohistochemical Subtypes Add Prognostic Value to Each Other in Breast Cancer Cohorts. Clin Cancer Res 2017; 23(24):7512–7520.

13. Lundberg A, Lindström LS, Li J et al. The long-term prognostic and predictive capacity of cyclin D1 gene amplification in 2305 breast tumours. Breast Cancer Res. 2019; 21(1):34.

14. Gown AM. Diagnostic Immunohistochemistry: What Can Go Wrong and How to Prevent It. Archives of Pathology & Laboratory Medicine 2016; 140(9):893–898.

